# Effectiveness of COVID-19 Vaccines Against SARS-CoV-2 Infection During a Delta Variant Epidemic Surge in Multnomah County, Oregon, July 2021

**DOI:** 10.1101/2021.08.30.21262446

**Authors:** Russell S. Barlow, Kevin Jian, Lindsey Larson

## Abstract

**Background:** Coronavirus disease 2019 (COVID-19) vaccines have been shown to be highly effective in preventing SARS-CoV-2 infection within controlled trials and real-world vaccine effectiveness (VE) studies. Recent reports have estimated reduced VE with the emergence and dissemination of the B.1.617.2 variant (“Delta variant”). We assess VE in Multnomah County, Oregon during a delta variant related epidemic expansion.

**Methods:** A test-negative design (TND) matched case-control analysis was performed to estimate the effectiveness of vaccination against SARS-CoV-2 infection during July 2021. Cases included a random sample of individuals that tested positive for SARS-CoV-2 and were reported by electronic laboratory report, were >15 years of age, and had no prior known SARS-CoV-2 infections. Controls were age and postal code matched individuals that tested negative for SARS-CoV-2 during July 2021. Immunization status was assessed using the Oregon ALERT Immunization Information system.

**Results:** 500 case-control pairs were matched (n=1000). 40.4% of cases were up-to-date on COVID-19 immunizations compared to 64.6% of controls. Effectiveness of any completed COVID-19 immunization was 73% (95% Confidence Interval [CI] 49-86%), 74% (95% CI 65-85%) for mRNA immunizations (BNT162b2, mRNA-1273), and 72% (95% CI 47-85%) for individuals partially immunized with mRNA immunizations.

**Conclusions:** Our findings estimate high, yet reduced, VE during Delta variant dissemination. These results highlight the importance of COVID-19 immunizations for reducing SARS-CoV-2 infection while juxtaposing the need for additional non-pharmaceutical interventions. Importantly, the reduced VE identified here may predict future reductions in vaccine performance in the context of ongoing viral genetic drift.

## Introduction

SARS-COV-2 has infected over 210 million individuals worldwide resulting in 4.4 million deaths as of August 17, 2021 [1]. Immunizations targeting the SARS-CoV-2 spike protein have been developed to prevent Coronavirus disease 2019 (COVID-19). In the United States (US), three immunizations have been granted Food and Drug Administration Emergency Use Authorization (FDA EUA) with Advisory Council on Immunization Practices (ACIP) recommendations including: Pfizer-BioNTech (BNT162b2) and Moderna (mRNA-1273) mRNA vaccines, and Janssen’s (Ad26.COV2.S) adenovirus vaccine [2-7]. Results from phase 3 randomized controlled trials of BNT162b2 and mRNA-1273 demonstrated high two-dose vaccine efficacy (∼95%) for preventing SARS-CoV-2 infection and COVID-19 with subsequent high vaccine efficacy at 6 month interim analyses [8-10]. Clinical trial results from Ad26.COV2.S demonstrated moderate single-dose efficacy (66.3%) in preventing COVID-19 [11]. Similarly, high vaccine effectiveness (VE) has been reported across several national and international observational studies for the mRNA vaccines[12-20].

Recent reports have indicated elevated vaccine breakthrough infections in the context of reduced prophylactic VE against the SARS-CoV-2 B.1.617.2 variant (“Delta variant”) with conflicting VE estimates ranging from 40%-88% [21-25]. Interpretation of these reports is complicated by differences in methods, study populations, care settings, immunization timelines, and possible selection and case ascertainment biases. To date, no studies have provided robust estimates of VE within a metropolitan population.

Multnomah County Health Department (MCHD) in Portland, Oregon monitors vaccine uptake and performance as part of our COVID-19 response. Previously, we evaluated baseline VE after immunization prevalence plateaued locally (65% of the total population immunized) in June, 2021 (unpublished supplemental data). Utilizing a test-negative design (TND) case-control approach, we estimated high VE consistent with published clinical trials and observational studies [12-20]. During July 2021, Multnomah County experienced a COVID-19 epidemic expansion likely driven by insufficient immunization coverage, increased social mixing, and the predominance of Delta variant (From 4% of sequenced cases during June to >75% during July 2021). This provided an opportunity to evaluate vaccine performance during Delta variant dissemination. Therefore, we performed a subsequent TND matched case-control assessment to provide instantaneous (point-in-time) countywide VE estimates during this epidemic expansion.

## Methods

### Assessment Population and Oregon’s COVID-19 Surveillance System

MCHD conducts active laboratory based surveillance of COVID-19 for 829,000 residents, who account for 19% of Oregon’s population [26]. Following ACIP guidance, Oregon began administering BNT162b2 and mRNA-1273 in December 2020 and subsequently Ad26.COV2.S in February 2021 to select populations [5-7]. Immunization eligibility was opened to all individuals >15 years of age during April 2021. By July 1st 2021, 70% of the county population over 15 years of age were estimated to have completed a COVID-19 immunization series with roughly 69% of those individuals receiving BNT162b2, 24% mRNA-1273, and 6% Ad26.COV2.S (unpublished data, [27]).

Physicians and laboratories are legally required to report all SARS-CoV-2 laboratory testing (positive, inconclusive, and negative) and clinically suspected cases of COVID-19 to local health departments via electronic laboratory reporting (ELR) or, in select settings, electronic case reporting (eCR), with demographic and clinical information. Data monitoring for consistency and completeness is performed on a routine basis including data linkage to the Oregon ALERT Immunizations Information System (ALERT IIS).

### SARS-CoV-2 Case and Control Definitions and Inclusion Criteria

To estimate COVID-19 vaccine performance against SARS-CoV-2 infection we utilized a TND matched case-control design including individuals that were tested for SARS-CoV-2 during July 2021 [28].

#### Cases

Primary SARS-CoV-2 infections among 15+ year olds that tested positive between July 1st and July 31st 2021 with an FDA EUA approved polymerase chain reaction (RT-PCR) or antigen test reported via ELR. Cases reported via eCR were not included for the following reasons: 1) eCR results are biased towards positive tests, 2) eCR testing primarily occurs within congregate settings and special populations where the prevalence of immunization, prevalence of testing, and prevalence of disease are often different from the general population (i.e. known or suspected effect modification).

#### Controls

ELR reported individuals >15 years of age that tested negative for SARS-CoV-2 with an FDA EUA approved RT-PCR or Antigen test between July 1st and July 31st 2021. If eligible controls were tested more than one time during the study period, they were only available for selection once.

### Vaccination Status Determination

Providers administering COVID-19 immunizations in Oregon are required to input immunizations into ALERT IIS within 72hrs of administration. Vaccination history was obtained by manually querying ALERT IIS for dates, type, and manufacturer for both cases and controls. Only provider-documented doses of vaccine were included in our assessment of vaccination status.

Vaccination status was determined according to the ACIP recommendations for all FDA EUA COVID-19 immunizations. For BNT162b2, 2 doses are required to be administered at least 21 days apart; for mRNA-1273, 2 doses are required to be administered at least 28 days apart; for Ad26.COV2.S a single dose was required [5-7]. To minimize differential exposure biases, the specimen date for the case was used to assess immunization status for both the case and the matched control. Shots were considered invalid if they were administered <14 days prior to the case’s positive test collection date.

Persons meeting the ACIP criteria for immunization were classified as up-to-date (abbreviated UTD). Persons who received prior valid COVID-19 immunizations (BNT162b2 and mRNA-1273 only) but failed to meet the ACIP criteria were classified as previously vaccinated, not up-to-date (abbreviated NUTD). Persons with no documented doses were classified as unvaccinated.

### Statistical Analyses

Both cases and controls were excluded from selection if they had documented prior RT-PCR or antigen confirmed SARS-CoV-2 infections. From the eligible cases, 500 were selected using simple random sampling. Eligible controls were stratified by age and postal code and were randomly matched 1:1 to cases using simple random sampling. Postal code was selected as a matching factor to help account for differences in disease, immunization,and testing prevalence, and unmeasured social factors. A sample size of 500 matched case and control pairs (n=1000) was estimated to provide >99% statistical power to detect an overall VE >50% with an immunization prevalence of at least 30%, or >80% statistical power to identify a VE >65% for specific vaccine types with a prevalence >5%.

Statistical assessment of differences between cases and controls were performed using the likelihood ratio chi-square test for categorical variables or the two sided Wilcoxon rank-sum test to assess differences between continuous characteristics, with a significance level of P<0.05 in both circumstances.

We assessed vaccination status, matching factors, and potential confounders using bivariate statistics. Conditional multivariate logistic regression analysis was performed to calculate odds ratios (OR) for the association between SARS-CoV-2 infection and receipt of FDA EUA COVID-19 immunizations between cases and controls. VE was estimated as (1-OR) X 100% [28]. VE estimates were assessed overall and individually for each FDA EUA vaccine. Conditional logistic regression was performed to preserve matching since vaccination status for controls was based on the specimen date of their matched case. Age and postal code were fixed in all models to control for residual or introduced confounding as a result of matching [29, 30].

This was considered public health practice by the MCHD Project Review Team as this assessment consisted only of analysis of routinely-collected reportable disease data. No cases or controls were contacted or interviewed as part of this assessment. All analyses were performed in SAS v 9.4 (SAS Institute Inc., Cary NC 2008).

## Results

### Case Population Characteristics

From July 1st through July 31st 2021, Multnomah County experienced an epidemic expansion coinciding with increased prevalence of Delta variant (32 of 41 [78%] sequences available at time of writing from July), resulting in 1398 confirmed SARS-CoV-2 infections being reported (Figure 1). Among these, 1156 (93.5%) met the inclusion criteria. During this same time, >30,000 negative lab results were reported for eligible controls. Cases and Controls were matched on age strata (ranked octiles of the case age distribution) and postal code, resulting in 500 matched pairs (500 cases, 500 controls). Immunization prevalence during July among individuals over 15 years of age ranged from 70-71% with increasing immunization prevalence with age (Table S1, [27]).

**Figure 1.**
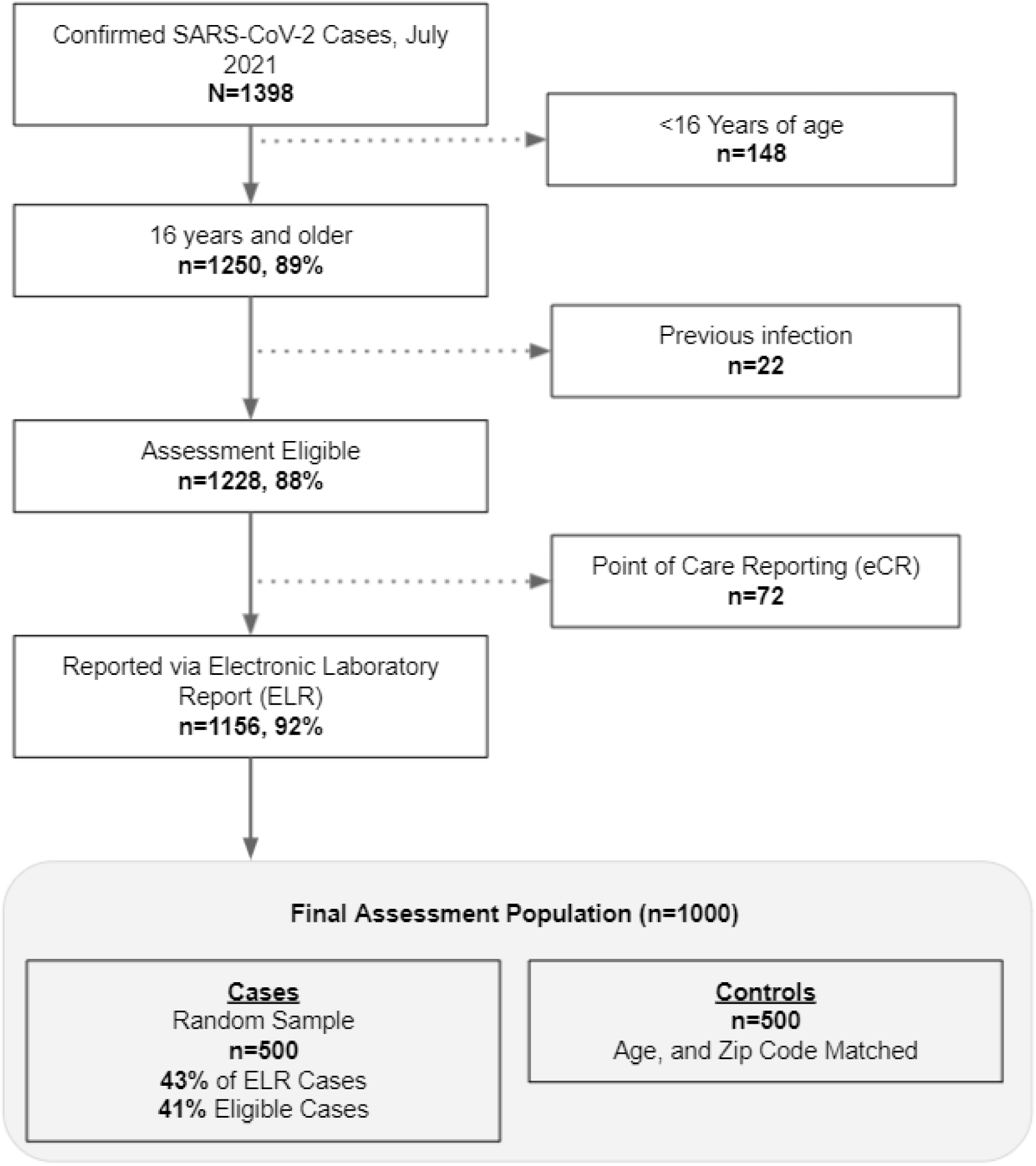
Case-Control Eligibility and Selection Flow chart, Multnomah County Oregon, July 2021.

The majority of cases were confirmed via RT-PCR (92%), median age was 37 years, 55% were female, 60% were white non-hispanic, 12% were of unknown race or ethnicity, 11% were black non-hispanic, 10 % were hispanic of any race, and 3.8% were asian non-hispanic (Table 1). Twenty-four (4.8%) cases were hospitalized as a result of COVID-19 and 1 death occurred. Forty percent of cases were fully immunized (UTD), 4% were previously immunized-NUTD, and 56% were completely unvaccinated.

**Table 1:**
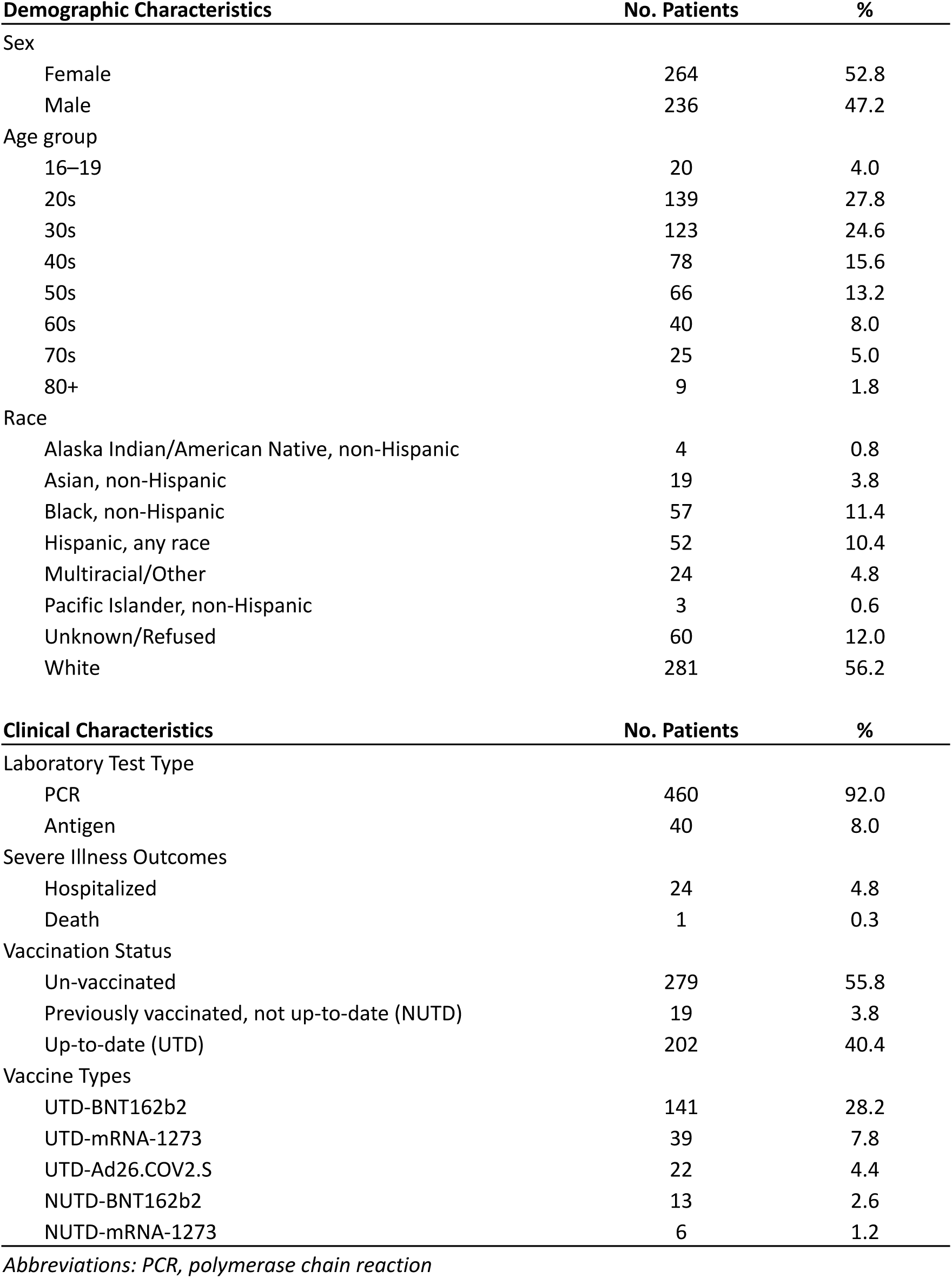
SARS-CoV-2 Case Demographic and Clinical Characteristics. Multnomah County Oregon, July 2021 (n=500)

| **Demographic Characteristics** | **No. Patients** | **%** |
| --- | --- | --- |
| Sex |  |  |
| Female | 264 | 52.8 |
| Male | 236 | 47.2 |
| Age group |  |  |
| 16–19 | 20 | 4.0 |
| 20s | 139 | 27.8 |
| 30s | 123 | 24.6 |
| 40s | 78 | 15.6 |
| 50s | 66 | 13.2 |
| 60s | 40 | 8.0 |
| 70s | 25 | 5.0 |
| 80+ | 9 | 1.8 |
| Race |  |  |
| Alaska Indian/American Native, non-Hispanic | 4 | 0.8 |
| Asian, non-Hispanic | 19 | 3.8 |
| Black, non-Hispanic | 57 | 11.4 |
| Hispanic, any race | 52 | 10.4 |
| Multiracial/Other | 24 | 4.8 |
| Pacific Islander, non-Hispanic | 3 | 0.6 |
| Unknown/Refused | 60 | 12.0 |
| White | 281 | 56.2 |
| **Clinical Characteristics** | **No. Patients** | **%** |
| Laboratory Test Type |  |  |
| PCR | 460 | 92.0 |
| Antigen | 40 | 8.0 |
| Severe Illness Outcomes |  |  |
| Hospitalized | 24 | 4.8 |
| Death | 1 | 0.3 |
| Vaccination Status |  |  |
| Un-vaccinated | 279 | 55.8 |
| Previously vaccinated, not up-to-date (NUTD) | 19 | 3.8 |
| Up-to-date (UTD) | 202 | 40.4 |
| Vaccine Types |  |  |
| UTD-BNT162b2 | 141 | 28.2 |
| UTD-mRNA-1273 | 39 | 7.8 |
| UTD-Ad26.COV2.S | 22 | 4.4 |
| NUTD-BNT162b2 | 13 | 2.6 |
| NUTD-mRNA-1273 | 6 | 1.2 |

*Abbreviations: PCR, polymerase chain reaction*

### Case and Control Comparisons

No differences between cases and controls were identified for sex or age. Cases were more likely to be black non-hispanic, compared to controls (Table 2). Cases were more likely than controls to be unvaccinated (55.8% [n=279] vs. 29.0% [n=145]) and were more likely to have immunization records in the ALERT IIS datasystem (96.6% [n=483] vs 94.0% [n=470], P=0.06). The median period from completion of COVID-19 immunizations until the matched case’s SARS-CoV-2 positive specimen were equivalent between cases and controls (102 days vs. 97 days P=0.29, respectively). No differences in this period were identified between cases and controls for specific COVID-19 Vaccines.

**Table 2:**
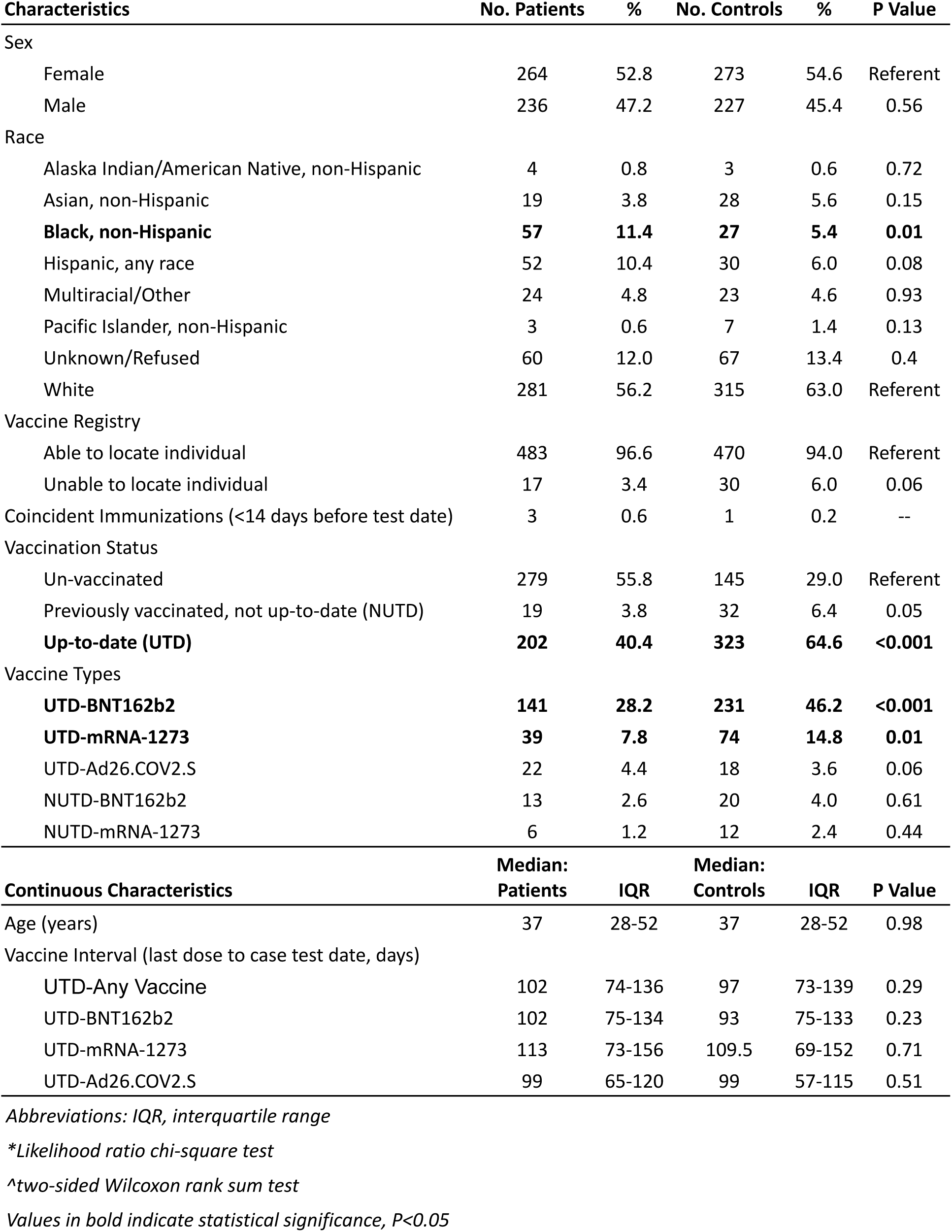
COVID-19 Case and Control Demographics and Immunization Comparisons. Multnomah County Oregon, July 2021 (n=1000)

| Characteristics | No. Patients | % | No. Controls | % | P Value |
| --- | --- | --- | --- | --- | --- |
| Sex |  |  |  |  |  |
| Female | 264 | 52.8 | 273 | 54.6 | Referent |
| Male | 236 | 47.2 | 227 | 45.4 | 0.56 |
| Race |  |  |  |  |  |
| Alaska Indian/American Native, non-Hispanic | 4 | 0.8 | 3 | 0.6 | 0.72 |
| Asian, non-Hispanic | 19 | 3.8 | 28 | 5.6 | 0.15 |
| **Black, non-Hispanic** | **57** | **11.4** | **27** | **5.4** | **0.01** |
| Hispanic, any race | 52 | 10.4 | 30 | 6.0 | 0.08 |
| Multiracial/Other | 24 | 4.8 | 23 | 4.6 | 0.93 |
| Pacific Islander, non-Hispanic | 3 | 0.6 | 7 | 1.4 | 0.13 |
| Unknown/Refused | 60 | 12.0 | 67 | 13.4 | 0.4 |
| White | 281 | 56.2 | 315 | 63.0 | Referent |
| Vaccine Registry |  |  |  |  |  |
| Able to locate individual | 483 | 96.6 | 470 | 94.0 | Referent |
| Unable to locate individual | 17 | 3.4 | 30 | 6.0 | 0.06 |
| Coincident Immunizations (<14 days before test date) | 3 | 0.6 | 1 | 0.2 | -- |
| Vaccination Status |  |  |  |  |  |
| Un-vaccinated | 279 | 55.8 | 145 | 29.0 | Referent |
| Previously vaccinated, not up-to-date (NUTD) | 19 | 3.8 | 32 | 6.4 | 0.05 |
| **Up-to-date (UTD)** | **202** | **40.4** | **323** | **64.6** | **<0.001** |
| Vaccine Types |  |  |  |  |  |
| **UTD-BNT162b2** | **141** | **28.2** | **231** | **46.2** | **<0.001** |
| **UTD-mRNA-1273** | **39** | **7.8** | **74** | **14.8** | **0.01** |
| UTD-Ad26.COVS2.S | 22 | 4.4 | 18 | 3.6 | 0.06 |
| NUTD-BNT162b2 | 13 | 2.6 | 20 | 4.0 | 0.61 |
| NUTD-mRNA-1273 | 6 | 1.2 | 12 | 2.4 | 0.44 |
| Continuous Characteristics | Median:<br>Patients | IQR | Median:<br>Controls | IQR | P Value |
| Age (years) | 37 | 28-52 | 37 | 28-52 | 0.98 |
| Vaccine Interval (last dose to case test date, days) |  |  |  |  |  |
| UTD-Any Vaccine | 102 | 74-136 | 97 | 73-139 | 0.29 |
| UTD-BNT162b2 | 102 | 75-134 | 93 | 75-133 | 0.23 |
| UTD-mRNA-1273 | 113 | 73-156 | 109.5 | 69-152 | 0.71 |
| UTD-Ad26.COVS2.S | 99 | 65-120 | 99 | 57-115 | 0.51 |

Abbreviations: IQR, interquartile range
\*Likelihood ratio chi-square test
^two-sided Wilcoxon rank sum test
Values in bold indicate statistical significance, $P < 0.05$

### Vaccine Effectiveness Assessment

Cases were significantly less likely to have started or completed all COVID-19 immunizations in bivariate analyses. In multivariate analyses, after adjustment for matching factors, we assessed additional potential confounding factors including race and ethnicity and sex. However, none of these factors were significantly different between cases and controls or confounded the point estimates (>10% change). The final model included immunization adjusted for age and postal code only (Table 3). Upon adjustment for matching factors, VE was moderately high at 73% for both UTD and NUTD vaccination statuses (Table 4).

**Table 3:** Case and Control Vaccination Status Associations With Odds of SARS-CoV-2 Infection. Multnomah County Oregon, July 2021 (n=1000)

| Characteristics | No.<br>Patients | % | No.<br>Controls | % | OR (95%CI) | Adjusted OR<br>(95% CI)* | P<br>Value |
| --- | --- | --- | --- | --- | --- | --- | --- |
| Vaccination Status |  |  |  |  |  |  |  |
| Un-vaccinated | 279 | 55.8 | 145 | 29.0 |  | Referent |  |
| **Previously vaccinated, not up-to-date (NUTD)** | **19** | **3.8** | **32** | **6.4** | **0.29 (0.16-0.54)** | **0.27 (0.14-0.51)** | **0.03** |
| **Up-to-date (UTD)** | **202** | **40.4** | **323** | **64.6** | **0.31 (0.23-0.41)** | **0.27 (0.19-0.37)** | **<0.001** |
| Vaccine Types |  |  |  |  |  |  |  |
| UTD-BNT162b2 | 141 | 28.2 | 231 | 46.2 | 0.30 (0.22-0.42) | -- | -- |
| UTD-mRNA-1273 | 39 | 7.8 | 74 | 14.8 | 0.26 (0.17-0.42) | -- | -- |
| UTD-Ad26.COV2.S | 22 | 4.4 | 18 | 3.6 | 0.57 (0.29-1.16) | -- | -- |
| NUTD-BNT162b2 | 13 | 2.6 | 20 | 4.0 | 0.29 (0.14-0.63) | -- | -- |
| NUTD-mRNA-1273 | 6 | 1.2 | 12 | 2.4 | 0.31 (0.11-0.86) | -- | -- |
| Aggregated Vaccination Status |  |  |  |  |  |  |  |
| NUTD-mRNA | 19 | 3.8 | 32 | 6.4 | 0.30 (0.16-0.55) | 0.28 (0.15-0.53) | 0.07 |
| **UTD-mRNA** | **180** | **36.0** | **305** | **61.0** | **0.29 (0.22-0.40)** | **0.26 (0.18-0.35)** | **<0.001** |
| UTD-Ad26.COV2.S | 22 | 4.4 | 18 | 3.6 | 0.57 (0.29-1.15) | 0.49 (0.24-1.02) | 0.63 |

Abbreviations: CI, confidence interval; OR, Odds Ratio; mRNA, either mRNA immunization (BNT162b2 or mRNA-1273)
\*matched and statistically adjusted by postal code and age
Values in bold indicate statistical significance, $P < 0.05$

**Table 4:** COVID-19 Vaccine Effectiveness Estimates. Multnomah County Oregon, July 2021 (n=1000)

| **Study Characteristics** | **Vaccine Effectiveness Estimate (%)\*** | **95% LL** | **95% UL** |
| --- | --- | --- | --- |
| **Vaccination Status** |  |  |  |
| Un-vaccinated |  | Referent |  |
| **Previously vaccinated, not up-to-date (NUTD)** | **73** | **49** | **86** |
| **Up-to-date (UTD)** | **73** | **63** | **81** |
| **Aggregated Vaccination Status** |  |  |  |
| NUTD-mRNA | 72 | 47 | 85 |
| **UTD-mRNA** | **74** | **65** | **82** |
| UTD-Ad26.COVS.S | 51 | -2 | 76 |

*Abbreviations: CI, confidence interval; LL, lower limit; UL, upper limit; mRNA, either mRNA immunization (BNT162b2 or mRNA-1273)*
*\*matched and statistically adjusted by postal code and age*
*Values in bold indicate statistical significance, $P < 0.05$*

Examination of specific immunizations revealed that both mRNA immunizations (BNT162b2 and mRNA-1273) had similar effect magnitudes and were not significantly different from each other (OR 0.3-0.26 for UTD and 0.29-0.31 for NUTD respectively). In contrast Ad26.COV2.S displayed a lower effect magnitude relative to the mRNA immunizations. Therefore, mRNA immunizations were combined into “mRNA UTD” and “mRNA NUTD” categories with a separate “UTD J&J” category to improve statistical power and investigate immunization specific VE in multivariate models.

After adjustment for matching factors, no other covariates were significantly different between cases and controls or confounded the association with immunization status. Cases were less likely than controls to have been UTD and NUTD for mRNA immunizations (OR 0.26 95% CI 0.15-0.35 P<0.001 and 0.28 9%CI 0.15-0.53 P=0.06 respectively, Table 3). In contrast, Ad26.COV2.S displayed reduced effect magnitude between cases and controls (OR 0.49 95%CI 0.24-1.02, P=0.60). VE for mRNA vaccines was 74% (95% CI 65-82%) for UTD individuals, 72% (95% CI 47-85%) for NUTD individuals, and 51% (95% CI -2-76%) for UTD-Ad26.COV2.S individuals (Table 4).

Exclusion of matched pairs where either a case or control was missing an immunization record (resultant n=910, 455 matched pairs), resulted in the same overall models (Table S2). As expected, the magnitude of the VE estimates increased across mRNA immunizations (79% 95%CI 70-86% for UTD and 78% 95%CI 57-89% for NUTD) as well as Ad26.COV2.S 58% (95%CI 12-80%) (Table S3). Sensitivity analyses varying acceptable dose spacing and the interval for protection following series completion (10-28 days), resulted in the same overall models with slight changes in the NUTD estimates only. Finally, utilization of unconditional vs. conditional logistic regression resulted in differences in VE estimation precision but did not change overall VE conclusions (data not shown).

## Discussion

To our knowledge, this is the first metropolitan county wide assessment of COVID-19 vaccine effectiveness during an epidemic expansion driven, in part, by the dissemination of Delta variant. We demonstrate instantaneous measures of VE for Pfizer-BioNTech (BNT162b2), Moderna (mRNA-1273), and Janssen (Ad26.COV2.S) vaccinations against SARS-CoV-2 infection during July, 2021. We enumerate that individuals >15 years of age with primary SARS-CoV-2 infection had highly reduced odds of receiving any COVID-19 immunizations compared to matched community controls, particularly among individuals that received mRNA immunizations. Our VE estimates indicate moderate to high protection during our local Delta variant driven epidemic expansion.

Our VE estimates for mRNA immunizations align with international and national observational studies that have estimated moderate to reduced VE for Delta variant (65-82% VE for mRNA immunizations, [21-23]). However, our findings do not support the dramatic reductions in VE reported elsewhere [24, 25]. Differences in study populations, vaccination types and vaccine uptake timelines, study design, and viral diversity may explain some of these differences. Despite methodological differences, our overall results align well with a VE assessment in New York State during Delta variant dissemination where vaccination availability and prevalence were similar [23]. Importantly, we were able to assess differences in vaccine performance across all COVID-19 immunizations including partially immunized populations. In contrast to other reports, we found no significant differences in vaccine performance between mRNA immunizations (BNT162b2, mRNA-1273) during the Delta variant driven epidemic expansion. However, we had insufficient statistical power to generate precise VE estimates for Ad26.COV2.S, though our point estimate indicated moderate VE.

Alarmingly, our VE estimates here are significantly reduced compared to a methodologically equivalent VE assessment we performed with cases tested a median of 6 weeks prior (June, 2021). During this previous assessment, immunization prevalence was equivalently static, disease prevalence was in decline, and the Delta variant did not represent a meaningful fraction of known COVID-19 cases in Multnomah County (Tables S4-S6). VE findings from our previous assessment aligned with clinical trial and observational study estimates of mRNA immunization performance [12-20]. Therefore, the reduction in VE reported here likely indicates that this change was not attributable to methodological differences with our approach. Interestingly, the interval from full vaccination until the matched case’s positive specimen date was equivalent between cases and controls. A longer interval for cases would have been indicative of waning of immunity with time. Collectively, these findings strongly suggest that the rapid reduction in VE within our population is explained by reduced overall vaccine performance against Delta variant [23].

The TND case-control methodology provided a strong instantaneous measure of vaccine performance during our Delta variant epidemic expansion. Our evaluation was semi-automated and interview-independent, facilitating repeatable VE assessments in near real-time. Additional advantages of this approach include being able to directly or indirectly control differences in age, health seeking behavior, case ascertainment, and social and demographic factors reported via ELR or related to postal code [28]. Nevertheless, this approach may limit generalizability to other populations where immunization uptake and Delta variant predominance differ. The immunization prevalence among controls was significantly lower than the locally estimated immunization prevalence during the study period (Table S1, [27]). This discrepancy is explained by non-homogenous vaccine prevalence across our population. Therefore, our case and control matching criteria help to account for immunization heterogeneity resulting in more robust VE estimates.

Exclusion of cases and possible controls that were reported via eCR testing minimized differential biases due to differences in reporting of test results and limited the influence of outbreaks within special populations. Consequently, our VE estimates may not be generalizable to congregate settings including long term care facilities where eCR testing predominates and vaccine performance may differ [31, 32]. Relatedly, we were unable to include additional factors associated with reduced vaccine performance including fragility and immune suppression, because medical record review was not performed [31-35].

The observational nature of this study has limitations. Namely, immunization ascertainment was marginally elevated for cases compared to controls because public health case investigators collect immunization records from cases and enter these data into ALERT IIS. This would differentially inflate immunization prevalence among cases resulting in VE underestimates. Similarly, provider documented, highly specific immunization status criteria reduced the possibility of misclassification. However, classification of cases and controls with missing immunization registry records as unvaccinated would also result in VE underestimates. Indeed, exclusion of matched pairs where either a case or a control was missing immunization status, resulted in significant increases in estimated VE. Surveillance data can result in biased case ascertainment if health seeking and COVID-19 testing differ with respect to disease severity or immunization status. This would tend to result in controls having lower immunization prevalence, biasing towards VE underestimates. Similarly, case ascertainment for prior SARS-CoV-2 infection likely resulted in differences in re-infection risk. This misclassification would tend to underestimate VE if more unvaccinated controls had protection due to primary infection [36, 37]. Finally, we only assessed VE for infection. Therefore, additional benefits of immunization including reduced COVID-19 severity are not realized with our methodology. Collectively, these small potential biases would serve to underestimate VE making our estimates conservative.

The moderate to high VE during a Delta variant related surge, highlights the overall success of COVID-19 immunizations in contributing to reductions in the SARS-CoV-2 infection rate at the population level.

Nonetheless, the high incidence of vaccine breakthrough infections within our population also illuminates the individual susceptibility to infection even among immunized individuals. Unfortunately, the high VE against SARS-CoV-2 infection reported here and elsewhere is likely insufficient to maintain an infection rate below 1 for variants as transmissible as Delta variant without additional non-pharmaceutical interventions (NPI) [38]. Thus, immunizations in concert with additional NPIs for both vaccinated and unvaccinated populations are necessary to limit epidemic oscillations. Future research should evaluate immunization benefits beyond infection including reducing severe outcomes [23, 37]. Studies to date have conflated the prophylactic benefits of immunization with reduced hospitalization and death for the estimation of VE [17, 19]. Whereas, declining prophylactic VE may result in increasing disease severity if the majority of protection is attributable to COVID-19 vaccines preventing infection. Therefore, research is needed to evaluate outcomes among vaccine breakthrough infections. Finally, booster doses have been proposed to improve VE [40]. However, boosting with the same formulation may not result in concomitant increases in VE if changes in protection are principally driven by viral genetic drift. Consequently, rational vaccine reformulations targeting multiple immunogenic components of the viral life cycle may be necessary.

## Data Availability

These data are protected by public health law and are not publicly available

## Acknowledgements

We thank Jennifer Vines and Kim Toevs (Multnomah County Health Department) for providing constructive input. We also thank Oregon ALERT IIS staff (Oregon Health Authority), Acute and Communicable Disease Interoperability Staff (Oregon Health Authority), and Multnomah County Information Technology and Orpheus/Opera “datamart” staff (Multnomah County Health Department) for developing and maintaining data structures utilized for this work.

## Supplementary Figures and Tables

**Table S1:**
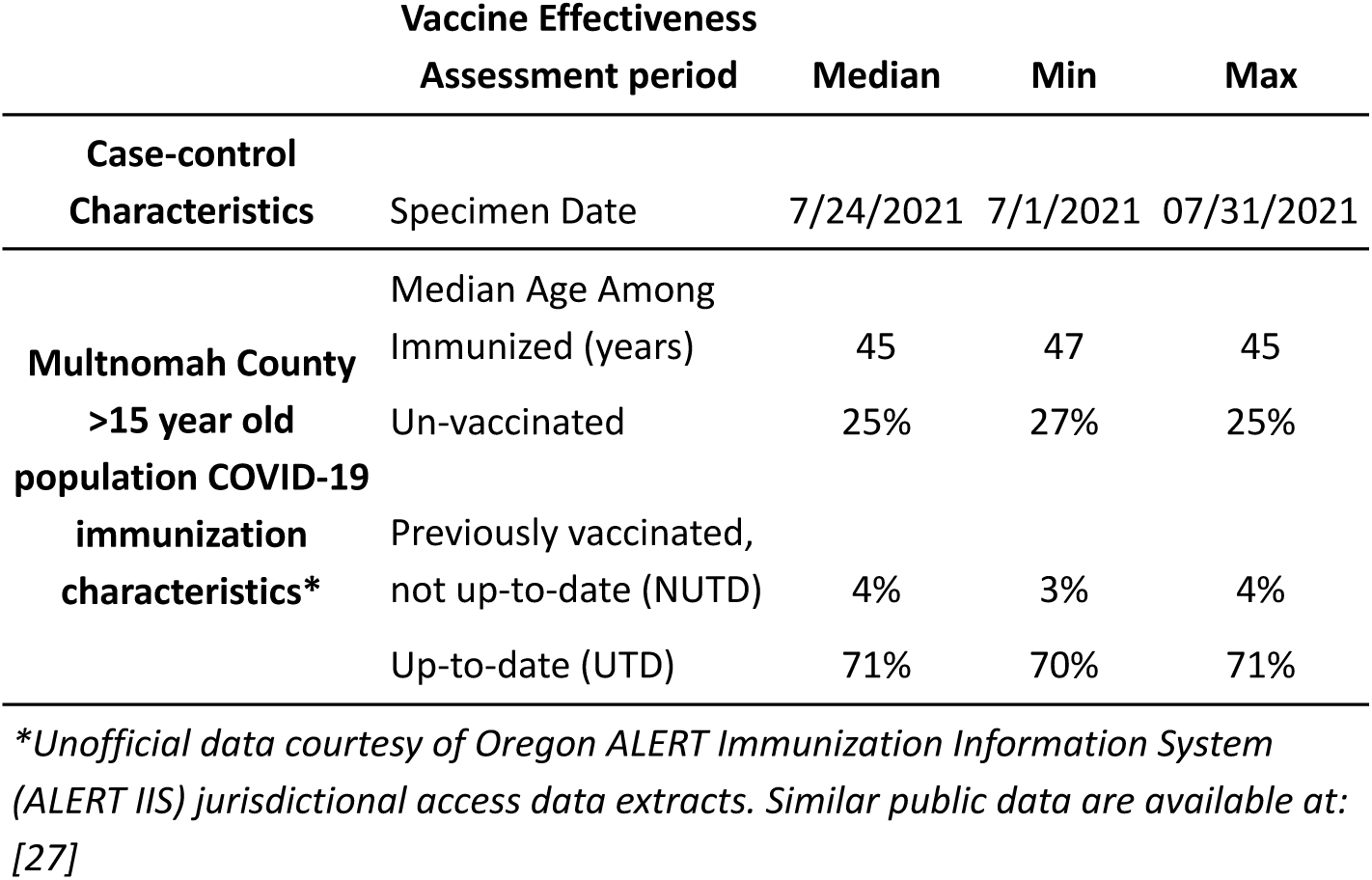
Population Immunization Prevalence Characteristics During Case-Control Vaccine Effectiveness Assessment, Multnomah County Oregon, July 2021.

**Table S2:**
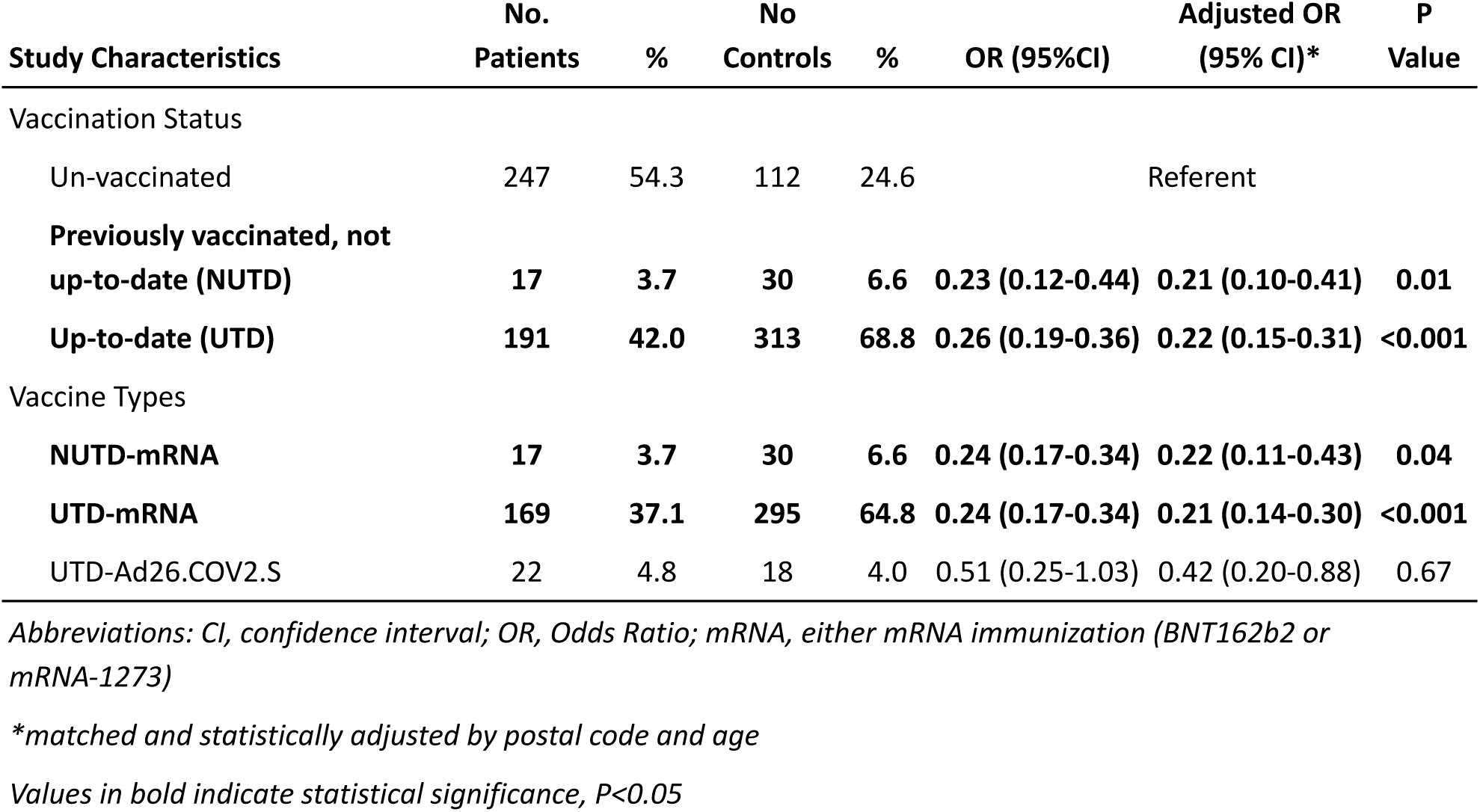
Case and Control Vaccination Status Associations With Odds of SARS-CoV-2 Infection, Excluding Case-Control Pairs With Missing Immunization Status. Multnomah County Oregon, July 2021 (n=910)

| Study Characteristics | No.<br>Patients | % | No<br>Controls | % | OR (95%CI) | Adjusted OR<br>(95% CI)* | P<br>Value |
| --- | --- | --- | --- | --- | --- | --- | --- |
| Vaccination Status |  |  |  |  |  |  |  |
| Un-vaccinated | 247 | 54.3 | 112 | 24.6 |  | Referent |  |
| **Previously vaccinated, not**<br>**up-to-date (NUTD)** | **17** | **3.7** | **30** | **6.6** | **0.23 (0.12-0.44)** | **0.21 (0.10-0.41)** | **0.01** |
| **Up-to-date (UTD)** | **191** | **42.0** | **313** | **68.8** | **0.26 (0.19-0.36)** | **0.22 (0.15-0.31)** | **<0.001** |
| Vaccine Types |  |  |  |  |  |  |  |
| **NUTD-mRNA** | **17** | **3.7** | **30** | **6.6** | **0.24 (0.17-0.34)** | **0.22 (0.11-0.43)** | **0.04** |
| **UTD-mRNA** | **169** | **37.1** | **295** | **64.8** | **0.24 (0.17-0.34)** | **0.21 (0.14-0.30)** | **<0.001** |
| UTD-Ad26.COV2.S | 22 | 4.8 | 18 | 4.0 | 0.51 (0.25-1.03) | 0.42 (0.20-0.88) | 0.67 |

Abbreviations: CI, confidence interval; OR, Odds Ratio; mRNA, either mRNA immunization (BNT162b2 or mRNA-1273)
\*matched and statistically adjusted by postal code and age
Values in bold indicate statistical significance, $P < 0.05$

**Table S3:** COVID-19 Vaccine Effectiveness Estimates, Excluding Case-Control Pairs With Missing Immunization Status. Multnomah County Oregon, July 2021 (n=910)

| Study Characteristics | Vaccine Effectiveness |  |  |
| --- | --- | --- | --- |
|  | Estimate (%)* | 95% CI LL | 95% CI UL |
| Vaccination Status |  |  |  |
| Un-vaccinated |  | Referent |  |
| **Previously vaccinated, not up-to-date (NUTD)** | **79** | **59** | **90** |
| **Up-to-date (UTD)** | **78** | **69** | **85** |
| Aggregated Vaccination Status |  |  |  |
| Un-vaccinated |  | Referent |  |
| **NUTD-mRNA** | **78** | **57** | **89** |
| **UTD-mRNA** | **79** | **70** | **86** |
| UTD-Ad26.COV2.S | 58 | 12 | 80 |

Abbreviations: CI, confidence interval; LL, lower limit; UL, upper limit; mRNA, either mRNA immunization (BNT162b2 or mRNA-1273)
\*matched and statistically adjusted by postal code and age
Values in bold indicate statistical significance, $P < 0.05$

**Table S4:** COVID-19 Case and Control Demographics and Immunization Comparisons. Multnomah County Oregon, June 2021 (n=600)

| Study Characteristics | No. Patients | % | No. Controls | % | P Value* |
| --- | --- | --- | --- | --- | --- |
| Sex |  |  |  |  |  |
| Female | 166 | 53.3 | 187 | 62.3 | Referent |
| Male | 134 | 44.7 | 113 | 37.7 | 0.08 |
| Race |  |  |  |  |  |
| Alaska Indian/American Native, non-Hispanic | 5 | 1.7 | 2 | 0.7 | 0.11 |
| **Asian, non-Hispanic** | **6** | **2.0** | **15** | **5.0** | **0.03** |
| **Black, non-Hispanic** | **42** | **14.1** | **24** | **8.0** | **0.04** |
| Hispanic, any race | 39 | 13.1 | 21 | 7.0 | 0.05 |
| **Multiracial/Other** | **9** | **3.0** | **22** | **7.3** | **0.03** |
| Pacific Islander, non-Hispanic | 2 | 0.7 | 4 | 1.3 | 0.31 |
| Unknown/Refused | 42 | 14.0 | 30 | 10.0 | 0.15 |
| White | 155 | 52.2 | 182 | 60.7 | Referent |
| Vaccine Registry |  |  |  |  |  |
| Able to locate individual | 279 | 93.0 | 286 | 95.3 | Referent |
| Unable to locate individual | 21 | 7.0 | 14 | 4.7 | 0.22 |
| Coincident Immunizations (<14 days before test date) | 7 | 2.3 | 8 | 2.7 | 1.00 |
| Vaccination Status |  |  |  |  |  |
| Un-vaccinated | 235 | 78.3 | 110 | 36.7 | Referent |
| Previously vaccinated, not up-to-date (NUTD) | 10 | 3.3 | 25 | 8.3 | 0.05 |
| **Up-to-date (UTD)** | **55** | **18.3** | **165** | **55.0** | **<0.001** |
| Vaccine Types |  |  |  |  |  |
| **UTD-BNT162b2** | **32** | **10.7** | **111** | **37.0** | **<0.001** |
| **UTD- mRNA-1273** | **13** | **4.33** | **50** | **16.7** | **0.01** |
| UTD-Ad26.COV2.S | 10 | 3.33 | 4 | 1.3 | 0.06 |
| NUTD-BNT162b2 | 8 | 2.7 | 15 | 5.0 | 0.61 |
| NUTD- mRNA-1273 | 2 | 0.67 | 10 | 3.3 | 0.44 |
| Continuous Characteristics | Median: Patients | IQR | Median: Controls | IQR | P Value^ |
| Age (Years) | 40 | 29-54 | 39.5 | 29-56 | 0.64 |
| Vaccine Interval (last dose to case test date, days) |  |  |  |  |  |
| UTD-BNT162b2 | 97.5 | 49.5-113.5 | 63 | 41-108 | 0.13 |
| UTD- mRNA-1273 | 88 | 58-144 | 83 | 33-121 | 0.07 |
| UTD-Ad26.COV2.S | 80.5 | 68-93 | 82 | 77-92 | 0.62 |

Abbreviations: IQR, interquartile range
\*Likelihood ratio chi-square test
^two-sided Wilcoxon rank sum test
Values in bold indicate statistical significance, $P < 0.05$

**Table S5:**
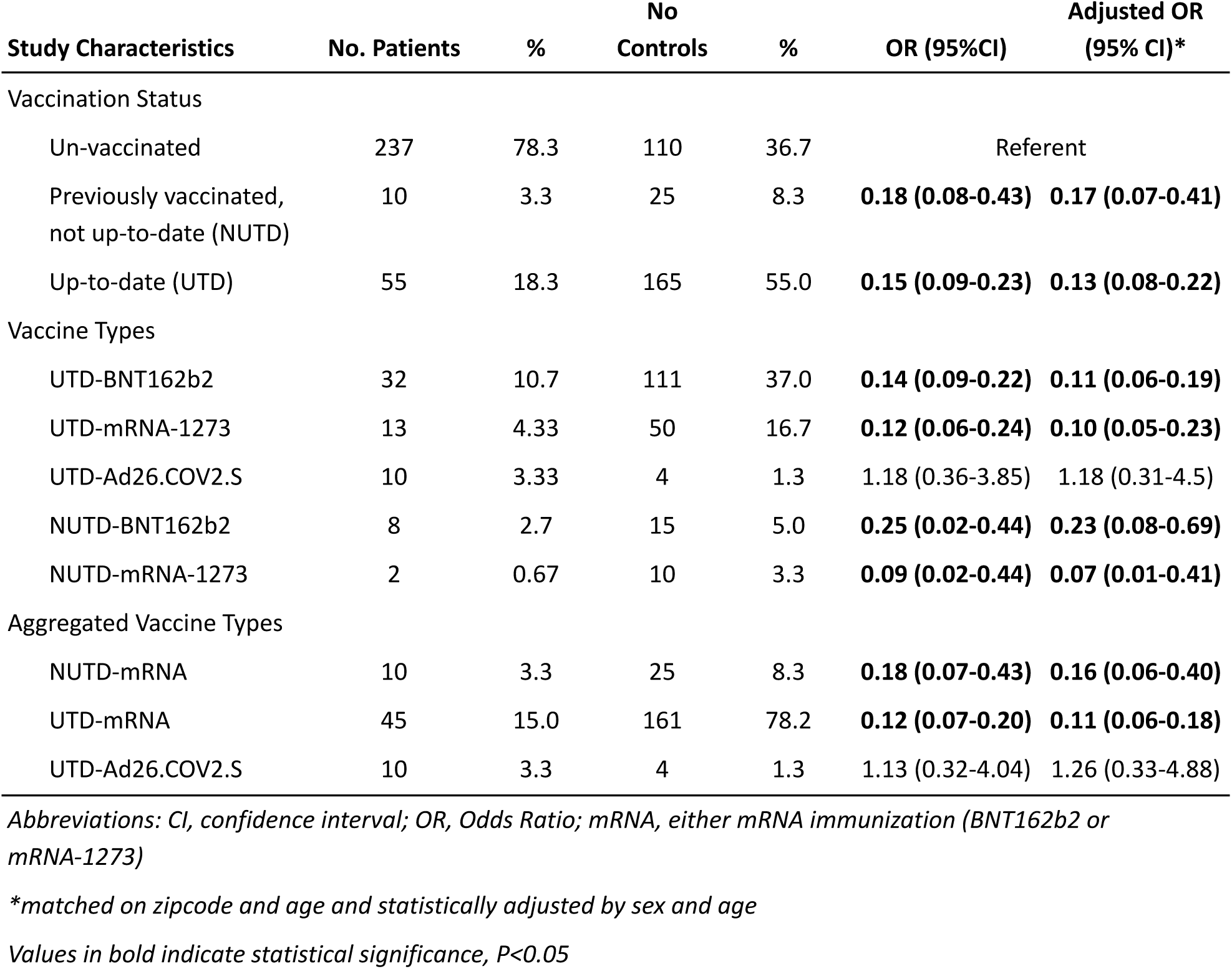
Case and Control Vaccination Status Associations With Odds of SARS-CoV-2 Infection. Multnomah County Oregon, June 2021 (n=600)

**Table S6:** COVID-19 Vaccine Effectiveness Estimates. Multnomah County Oregon, June 2021 (n=600)

| Study Characteristics | Vaccine Effectiveness |  |  |
| --- | --- | --- | --- |
|  | Estimate (%)* | 95% LL | 95% UL |
| Vaccination Status |  |  |  |
| Un-vaccinated |  | Referent |  |
| Previously vaccinated, not up-to-date (NUTD) | **83** | **59** | **93** |
| Up-to-date (UTD) | **87** | **78** | **92** |
| Aggregated Vaccination Status |  |  |  |
| NUTD-mRNA | **84** | **60** | **94** |
| UTD-mRNA | **89** | **82** | **94** |
| UTD-Ad26.COV2.S | -26 | -488 | 67 |

*Abbreviations: CI, confidence interval; LL, lower limit; UL, upper limit; mRNA. either mRNA immunization (BNT162b2 or mRNA-1273)*
*\*matched on zipcode and age and statistically adjusted by sex and age*
*Values in bold indicate statistical significance, $P < 0.05$*

